# Simulating Ultrasound images from CT Scans

**DOI:** 10.1101/2023.01.16.23284615

**Authors:** Sahar Almahfouz Nasser, Amit Sethi

## Abstract

Anatomical information in ultrasound (US) imaging has not been exploited fully because its wave interference pattern (WIP) has been viewed as speckle noise. We tested the idea that more information can be retrieved by disentangling the WIP rather than discarding it as noise. We numerically solved the forward model of generating US images from computed tomography (CT) images by solving wave-equations using the Stride library. By doing so, we have paved the way for using deep neural networks to be trained on the data generated by the forward model to simulate the solution of the inverse problem, which is generating the CT-style and CT-quality images from a real US image. We demonstrate qualitative features of the generated images that are rich in anatomical details and realism.

## 1 INTRODUCTION

### 1.1 Background

Ultrasound is a non-ionizing imaging modality that makes it a vital tool for medical imaging and image-guided interventions. It is also portable and realtime, unlike other imaging modalities, such as magnetic resonance imaging (MRI) and computed tomography (CT), which are rich in detail but are bulky, unweidly and not real time. However, the presence of speckle noise-like artifacts, blurring, and shading issues reduce the diagnostic value of US as an imaging modality.

Developing methods for US denoising is essential to conduct a better diagnosis, assessment, and image-guided interventions in real time (Duarte-Salazar et al., 2020). The main artifact in US is often said to be speckle noise. Speckle noise is a granular noise with a multiplicative nature (Wagner, 1983), and (Kaplan and Ma, 1994). For instance, In synthetic-aperture radar (SAR) images the observed signal, can be described according to (Mather and Tso, 2016) as follows:

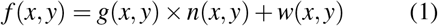

where *f* (*x, y*) is the observed signal, *g*(*x, y*) is the original signal, *n*(*x, y*) is a multiplicative noise, and *w*(*x, y*) is an additive noise.

However, while the noise in US appears to be speckled in nature, it is actually wave interference pattern (WIP), which is produced by additive and destructive interference of the ultrasound waves with the tissue – a phenomenon which is known as scattering. There are two types of scattering: diffuse scattering and coherent scattering. Diffuse scattering generates speckles in the image, while the coherent one yields clear, dark, and bright features. Speckle noise in US is, therefore, a signal-dependent noise, which relies on the structure and the imaging factors of the imaging system (Singh et al., 2017).

In this work, we describe a simulation method for US images starting from 2D CT images. The main purpose of this simulation is to generate paired images to learn the inverse model from US to CT, so that deep neural networks can be trained for real time and portable simulation of CT-like images with rich anatomical details from regular real time US images and videos. Such a simulation will bring the best of the two modalities – portability and real time nature of US with the clarity and details of CT – to diagnostics and surgical intervention. Surprisingly, even the forward model to simulate US from CT had not been fully described in one place, and nor made available as a software before our work.

In the rest of the paper, we describe US physics, related work, and our proposed method. Then, we show qualitative results of our method and conclude with possible directions for future work.

### 1.2 Ultrasound Physics

The US is a non-ionizing type of energy, which makes it suitable for real-time and interactive medical imaging.

US generation is based on the reverse piezoelectric effect while detecting it is based on the piezoelectric effect. The US waves propagate in the tissue in two ways longitudinal and transverse. In longitudinal propagation, the wave propagates in the same direction as the perturbation causing it. However, if the wave propagation is perpendicular to the disturbance generating it, it is called transverse propagation.

US interacts with tissues in four ways: reflection, refraction, absorption, and scattering (Tole et al., 2005).

#### 1.2.1 Reflection

Reflection happens at the boundaries between adjacent tissues – the acoustic boundaries. Based on the size of the boundary relative to the US beam wavelength, or the irregularities of the surface of the reflector, we can divide the reflection into two categories: the specular reflection and the non-specular reflection. Specular reflection happens when the boundary is smooth and longer than the beam dimensions, while non-specular reflection occurs when the size of the reflector is smaller than the wavelength of the ultrasound beam.

The reflection coefficient on the acoustic surface is given by

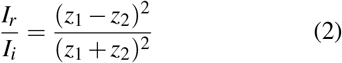

where *I*_*i*_ is the intensity of the incident beam, *I*_*r*_ is the intensity of the reflected beam, *z*_1_ is the acoustic impedance of the first medium, and *z*_2_ is the acoustic impedance of the second medium.

The more the difference between the impedance values (acoustic mismatch), the larger the echo.

The irregularity in the shape of the reflecting surface or its small dimensions reflects the incident beam in many directions is known as US scattering.

The scattering strongly depends on the US frequency, so it increases as the frequency increases.

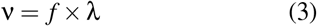

where ν is the sound velocity, f is the frequency, and λ is the wavelength. See Figure 1.

**Figure 1:**
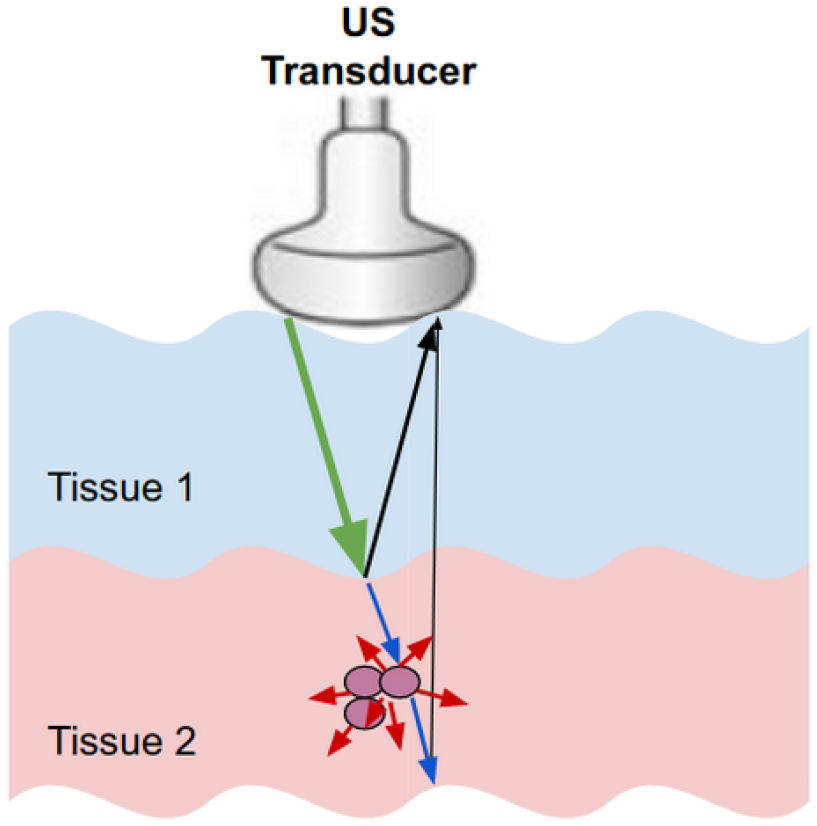
Ultrasound interaction with tissues. The green, black, red, blue arrows represent the incident wave, the specular reflection, the non-specular reflection, and the refracted wave, respectively.

#### 1.2.2 Absorption

During absorption, the US energy gets converted into heat. Three factors affect absorption – the viscosity of the medium, the relaxation time of the medium, and the frequency of the beam. Absorption increases in direct proportion to all three factors. The viscosity is generated from the frictional forces between the particles. The relaxation time is the duration required for the particles of the medium to get back to their mean position after getting displaced by the US waves. Absorption also increases with the beam frequency, although increased frequency can enhance details in the US image.

#### 1.2.3 Attenuation

Attenuation is the reduction of the beam intensity caused by the total losses throughout the propagation. Based on the power law (Cong et al., 2013), the energy attenuation of the ultrasound wave after its propagation in the medium for a distance d is given by:

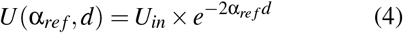

where α is an attenuation parameter related to the properties of the propagation medium.

### 2 RELATED WORK

As we mentioned in the abstract, many researchers tried to simulate ultrasound images simply by modeling US with speckle noise and ignoring other interactions of ultrasound with the tissue.

Goodman modeled speckle noise in laser images by a Rayleigh distribution (Goodman, 1975). Wagner et al (Wagner, 1983) represented the speckle noise by a Rician model. While Shankar (Shankar, 2000) came up with Nakagami distribution to describe speckle noise. Usually, Gamma distribution is the best approximation of speckle noise in SAR images (Ayed et al., 2005). Zimmer (Zimmer et al., 2000) modeled speckle noise in ultrasound liver images by a lognormal distribution. Tao et al in (Tao et al., 2006) proved that Gamma and Weibull distributions are better approximations of speckle noise in clinical cardiac ultrasound images than normal or log-normal distributions.

In (Achim et al., 2001) and (Rabbani et al., 2008), the authors proposed a method to convert the multiplicative noise into an additive noise by logarithmically transforming the image as follows:

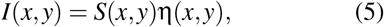

where *I* is the noisy observation (the US image), S is the noise-free image, and η represents the multiplicative speckle noise.

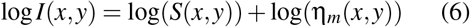

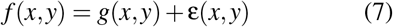

Shams et al (Shams et al., 2008) proposed a novel method for simulating ultrasound images from 3D CT scans. In the proposed method, the authors started with edge detection of the CT image to calculate the reflection coefficients. Then they generated the scattering image using FieldII (Jensen, 1996) by placing scatterers with strength randomly chosen by Field II from a normal distribution. The authors indicated that using this method to create a realistic speckle pattern is very computationally expensive. For instance, simulating a B-mode image with 128 RF scan lines takes nearly two days.

Kutter et al (Kutter et al., 2009) proposed a simulation-based registration pipeline of US to CT images in real-time. They developed a simple ray-based modeling of ultrasound images using OpenGL software (woo,). They used a Lambertian scattering model to simulate the scattered signal. And they generated a scattering image using Field II software. In (Reichl et al., 2009), the US intensity at the location of the probe was adjusted at first. Then, the amount of intensity transmitted, reflected, or absorbed along each column (each scanline) of the image was computed for every pixel according to the propagation characteristics. After that, the reflection and the absorption were subtracted from the incident intensity at every pixel. Finally, in the post-processing stage, artifacts such as speckle noise and blurring were added to the ultrasound images. In their proposed work, speckle noise was characterized by a Rayleigh distribution.

Feng Gu et al. proposed a genrative adversarial network (GAN) to model the speckle noise in synthetic aperture radar (SAR) images (Gu et al., 2019).

In this work, we present a novel method for simulating the interaction pattern between the ultrasound waves and the tissue, that is inspired by the underlying physics of ultrasound image generation, starting from CT images of different body parts.

## 3 PROPOSED METHOD

Our proposed method for generating ultrasound images from CT images consists of two stages – generating speed of sound images from CT images, and generating US images from speed of sound images. The code of our proposed method is available at (Almahfouz Nasser and Sethi,).

### 3.1 Generating speed of sound images from CT images

To generate the speed of sound images from CT images, we use the fact that the intensity value of a specific pixel of a 2D CT image represents the Hounsfield unit (HU) of the underlying tissue that corresponds to that pixel. HU is a measure of X-ray attenuation in the tissue. Given a tissue *x*, the HU is given by:

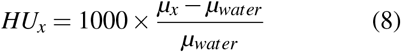

where *µ*_*x*_ is the total linear attenuation coefficient of the tissue *x* at a given x-ray energy. *µ*_*x*_ of a tissue x can be computed from multiplying the mass density of a tissue x (ρ_*x*_) by the weighted sum of the mass attenuation coefficients of all the elements which compose the tissue x, as shown in the following equation:

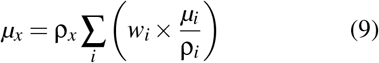

Where 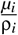 is the mass attenuation coefficient of the el-ement (i) in cm^2^/g. Table 1 shows the elemental composition (*w*_*i*_ values) of a few of the body tissues taken from the ITIS database (ITI,).

**Table 1:** Examples of the composition of a few of the body tissues. This table does not include the values of all the elements of the tissue composition, other elements, such as silicon and phosphorus, can be found in (Ele,).

| Tissue | Hydrogen | Carbon | Nitrogen | Oxygen | Sodium | Magnesium | Sulfur | Chlorine | Argon | Potassium |
| --- | --- | --- | --- | --- | --- | --- | --- | --- | --- | --- |
| Air | 0 | 0.00015 | 0.78 | 0.21 | 0 | 0 | 0 | 0 | 0.0047 | 0 |
| Liver | 0.63 | 0.073 | 0.013 | 0.28 | 0.00054 | 0 | 0 | 0.0006 | 0.00058 | 0.00035 |
| Prostate | 0.64 | 0.046 | 0.011 | 0.3 | 0.00054 | 0.0002 | 0.00039 | 0 | 0 | 0.00032 |
| Fat | 0.5 | 0.37 | 0.003 | 0.13 | 0.00018 | 0.0005 | 0.00013 | 0.00012 | 0 | 0 |
| Kidney | 0.63 | 0.069 | 0.013 | 0.28 | 0.00054 | 0.0004 | 0.00039 | 0.00035 | 0 | 0.00032 |
| Urinary Bladder Wall | 0.64 | 0.049 | 0.011 | 0.29 | 0.00054 | 0.0004 | 0.00039 | 0.00052 | 0 | 0.00047 |
| Urine | 0.66 | 0.0025 | 0.0043 | 0.33 | 0.0011 | 0.0002 | 0 | 0.001 | 0 | 0.00031 |
| Water | 0.67 | 0 | 0 | 0.33 | 0 | 0 | 0 | 0 | 0 | 0 |

Given the kilovoltage peak applied to the X-ray tube for capturing CT images is 120 kPv, and *w*_*i*_ values from (ITI,), we can compute the mass attenuation coefficient of each element in the elemental composition of a certain tissue by using NIST software (NIS,). NIST allows us to compute the mass attenuation coefficients of the tissues, which, in turn, allows us to compute their corresponding HUs by substituting the values in equation 8.

Now having the speed of sound values and the corresponding HU values of the tissues at 37 Celsius, we can generate the speed of sound images from the corresponding CT images.

We simulate the attenuation of US waves in tissues using equation (4),

### 3.2 Generating US images from speed of sound images

In the second stage of our proposed method, we simulate the US images from the speed of the sound images. For simulating ultrasound images from the speed of sound images which we generate, we use Stride software (Cueto et al., 2022). Stride is an open-source library for ultrasound computed tomography. Unlike the methods based on full-waveform inversion, Stride is not computationally expensive. Stride is user-friendly software, and the code can be run on CPUs and GPUs. This tool is based on a domain-specific language called Devito which generates solvers of the wave-equation.

We can summarize the overall workflow of this software to reconstruct the image of the tissue from the measurements as follows:

1. The sensors produce acoustic waves, which propagate throughout the medium. These propagated waves get reflected on the acoustic boundaries of the medium.
2. The reflected waves get captured by the receivers.
3. The acquired data is used to reconstruct the physical properties of the medium, for instance, the speed of the wave through it and its density.
4. The reconstruction procedure minimizes the misfit between the recorded measurements and the numerically modeled ultrasound data.

Thus from a set of measurements of the pressure wave field *u*, we can build an accurate model of the discrete wave velocity *C* (or 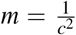) by consider-ing it as a partial differential equation-constrained optimization problem, where the objective function is given by:

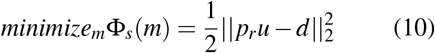

with 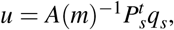, where *p*_*r*_ is the sampling operator of receiver locations, 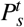 represents the injection operator at source locations, *A*(*m*) is the discrete isotropic wave equation matrix, *u* is the discrete pressure wave field, *q*_*s*_ is the pressure source, and *d* is the measured date.

By solving the optimization problem based on the gradient method (Plessix, 2006) (Haber et al., 2012) we get:

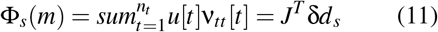

where *n*_*t*_ is the number of steps, δ*d*_*s*_ = *p*_*r*_*u − d* is the data residual between the measured and the modeled data, *J* is the Jacobian operator, ν_*tt*_ is the second-order time derivative of the adjoint wave field 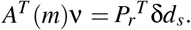

## 4 RESULTS AND DISCUSSION

We designed a curvilinear transducer similar to the one used by clinicians for abdominal imaging. The transducer has a central frequency of 2.5 MHZ, a number of elements equals 64, a radius of the curvature equals 5R, and a curvature equals 50 mm.

The simulation consists of a forward pass and a backward pass. For the backward pass, rather than starting from a fixed speed of sound, we started from an initial estimation of the speed of the sound image to improve convergence. This initial estimation is a blurred version of the ground truth speed of the sound image. See Figure 2.

**Figure 2:**
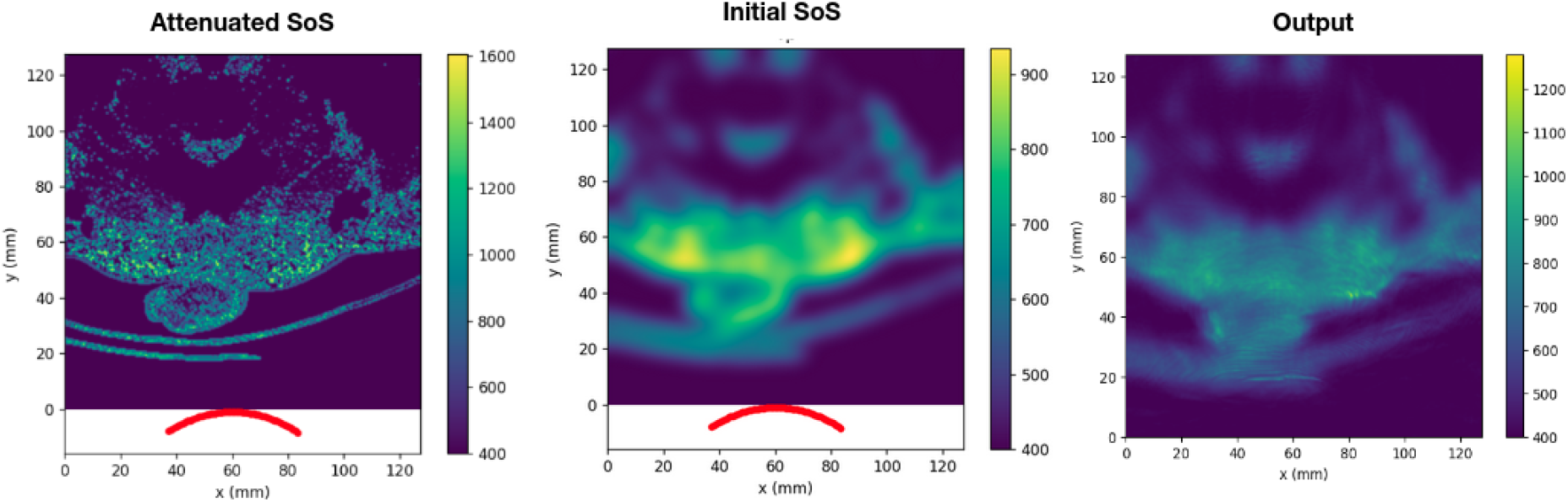
The images from left to right are the attenuated speed of sound (SoS) image (the input of the forward pass), the initial speed of sound image (the input of the backward pass), and the output of Stride software.

Figure 3 shows a few results of our proposed method for ultrasound simulation. One can see in the simulated US image the arc artifacts that are found in real ultrasound images when a curvilinear transducer is used.

**Figure 3:**
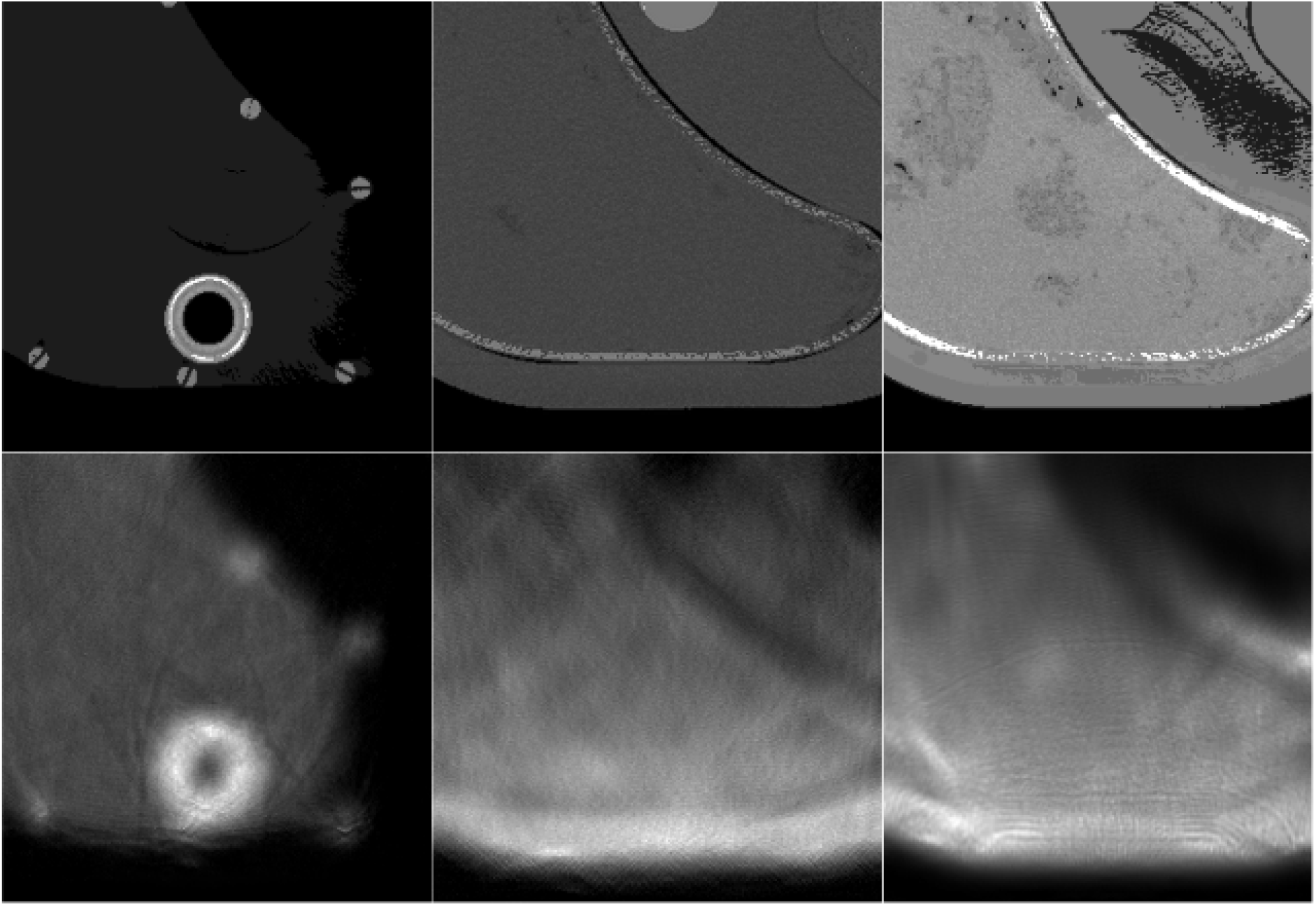
A few example output images of our proposed method for ultrasound simulation. The images are three different slices of a phantom of the liver. The first row contains the speed of sound images generated from the corresponding CT images (Ima,). The second row contains the simulated ultrasound images. The experiments from left to right are a simulation using a linear probe without attenuation, a simulation using a linear probe with attenuation, and a simulation using a curvilinear probe with attenuation correspondingly.

In figure 4, we try to qualitatively assess our results by visually comparing the simulated images with real US images. In figure 4, the CT images and real US images were taken from (Ima,), (AbU,).

**Figure 4:**
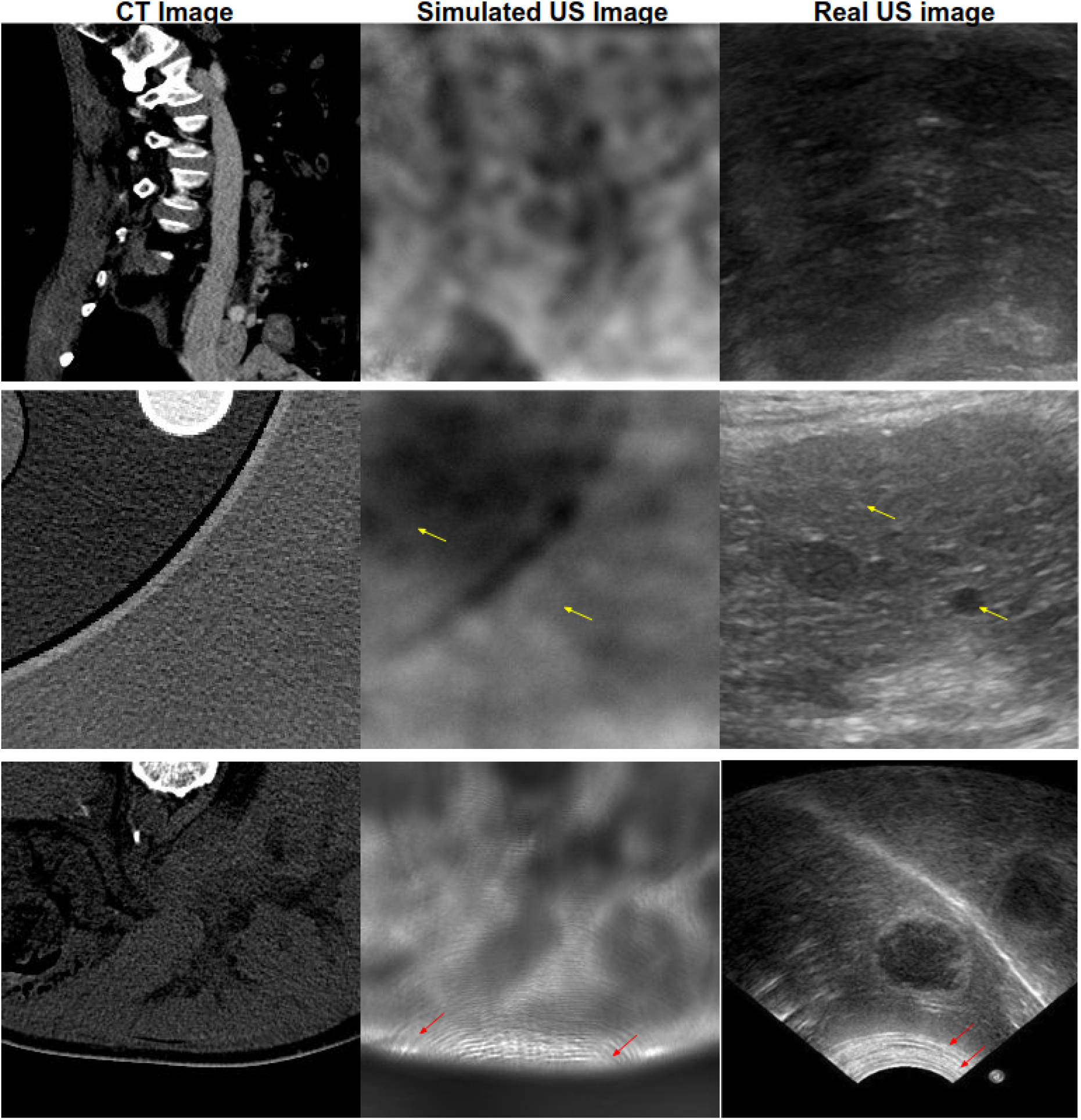
The first two columns contain CT images and the corresponding simulated ultrasound images, while the third column contains real ultrasound images which do not correspond to CT images. The red arrows indicate the arc artifacts in both real and simulated US images. The yellow arrows indicate the granular texture in simulated and real US images. For a precise comparison between our simulated images and real US ones, we are planning further experiments in which we acquire CT scans of a phantom to simulate ultrasound images using our proposed method. After that, we can compare the simulated US images with the real US images of the same phantom.

We refer to the work presented in (Kutter et al., 2009) and (Reichl et al., 2009) for visual comparison with the results of our proposed method. In our simulation, we use Stride software built upon the open-source programming language (Python). While methods (Kutter et al., 2009) and (Reichl et al., 2009) use a Matlab-based software called Field II. For a fair comparison between our methods and other methods, our extended paper should incorporate a comparison between these methods for the same ground truth CT images, and the same US imaging probe.

## 5 CONCLUSIONS

In this work, we showed promising results in simulating US images from CT images. The wave interference pattern and other artifacts were similar to real US images. Our proposed method for obtaining the simulated ultrasound images from the CT images needs further improvement. After optimizing the hyper-parameters of our simulation we will be able to form a paired dataset (CT-Ultrasound pairs) which can be used for training a generative adversarial network GAN in a supervised manner and finally testing it on reconstructing the CT images from the real US images. The potential utility of this work is to train deep neural networks for the inverse problem of simulating CT images from the given US images, which can aid clinicians in diagnosis and surgical intervention.

## Data Availability

All data produced are available online at:
Cancer imaging archive.
https://www.cancerimagingarchive.net/. Accessed:
2021-10-24.
Computing the mass attenuation coefficients of
the elemental composition of the tissues.
https://www.nist.gov/pml/xcom-photon-cross-
sections-database. Accessed: 2021-10-24.
Element composition of the body tissues.
https://itis.swiss/virtual-population/tissue-
properties/database/elements/. Accessed: 2021-
10-24.
ITIS Foundation. https://itis.swiss/virtual-
population/tissue-properties/database/elements/.
Accessed: 2021-10-24

https://www.nist.gov/pml/xcom-photon-cross-sections-database

https://itis.swiss/virtual-population/tissue-properties/database/elements

## ACKNOWLEDGEMENT

This work would not have been possible without the financial support of the Qualcomm Innovation Fellowship Award, India. We are indebted to the developer of Stride, Mr. Carlos Cueto from Imperial College London, for his feedback and support.

## Notes

### Funding Statement

This study was funded by the Qualcomm Innovation Fellowship Award, India.

### Author Declarations

The study used (or will use) ONLY openly available human data that were originally located at: Cancer imaging archive. https://www.cancerimagingarchive.net/. Accessed: 2021-10-24. Computing the mass attenuation coefficients of the elemental composition of the tissues. https://www.nist.gov/pml/xcom-photon-cross- sections-database. Accessed: 2021-10-24. Element composition of the body tissues. https://itis.swiss/virtual-population/tissue- properties/database/elements/. Accessed: 2021- 10-24. ITIS Foundation. https://itis.swiss/virtual- population/tissue-properties/database/elements/. Accessed: 2021-10-24

